# An AI-driven machine learning approach identifies risk factors associated with 30-day mortality following total aortic arch replacement combined with stent elephant implantation

**DOI:** 10.1101/2025.03.06.25323548

**Authors:** Shuai Zhang, Lulu Li, Jingyu Wang, Yuan Li, Cuntao Yu, Xiaogang Sun, Jing Sun, Xiangyang Qian

**Author notes:** Corresponds to: Xiangyang Qian. Shuai Zhang and Lulu Li made equal contributions to this project and should be regarded as co-primary authors.

## Abstract

**Objectives:** During emergency surgery, patients with acute type A aortic dissection (ATAAD) experience unfavorable outcomes throughout their hospital stay. The combination of total aortic arch replacement (TAR) and frozen elephant trunk (FET) implantation has become a dependable choice for surgical treatment. The objective of this research was to utilize a machine learning technique based on artificial intelligence to detect the factors that increase the risk of mortality within 30 days after surgery in patients who undergo TAR in combination with FET.

**Methods:** From January 2015 to December 2020, a total of 640 patients with ATAAD who underwent TAR and FET were included in this study. The subjects were divided into a test group and a validation group in a random manner, with a ratio of 7 to 3. The objective of our research was to create predictive models by employing different supervised machine learning techniques, such as XGBoost, logistic regression, support vector machine (SVM), and random forest (RF), to assess and compare their respective performances. Furthermore, we employed SHapley Additive exPlanation (SHAP) measures to allocate interpretive attributional values.

**Results:** Among all the patients, 37 (5.78%) experienced perioperative mortality. Subsequently, a total 50 of 10 highly associated variables were selected for model construction. By implementing the new method, the AUC value significantly improved from 0.6981 using the XGBoost model to 0.8687 with the PSO-ELM-FLXGBoost model.

**Conclusion:** In this study, machine learning methods were successfully established to predict ATAAD perioperative mortality, enabling the optimization of postoperative treatment strategies to minimize the postoperative complications following cardiac surgeries.

## 1. Introduction

Acute Type A Aortic Dissection (ATAAD) is a severe condition linked to significant fatality rates (16−19% for surgical mortality)^1–2^. The mortality of ATAAD increases over time, with a rate of 1% per hour^3^. ATAAD is primarily treated with Total Aortic Arch Replacement (TAR) using deep hypothermic circulatory arrest. The approaches to surgically treating ATAAD lesions differ because of the complex and diverse characteristics of these lesions^4^. In contrast to Western nations, the most common surgery for ATAAD in China is TAR with the frozen elephant trunk (FET) method^5^.

Nevertheless, despite the progress made in surgical methods and perioperative care, the fatality rate after surgical intervention remains elevated. Given its intricate nature and elevated risk, the success of TAR+FET largely depends on the patient’s condition before surgery and the specifics of the surgical process. Premature death is linked to numerous high-risk elements such as advanced age, inadequate pre-surgery organ perfusion, heart tamponade, low blood pressure, stroke after surgery, continuous renal replacement therapy, low cardiac output syndrome, and multiple organ dysfunction^6^. Hence, it is crucial to create suitable forecasting models for evaluating linked hazards and recognizing populations at high risk in clinical settings.

Logistic regression is widely applied in the construction of various medical predictive models, serving as a linear model for solving binary classification problems. However, Logistic regression has several limitations, such as assuming a linear relationship between independent and dependent variables, sensitivity to multicollinearity, and challenges with imbalanced categories. Consequently, this study applies an improved machine learning method called RF-PSO-FLXGBoost to establish an accurate prediction model.

## 2. Methods

### 2.1. Study population and Definition

From January 2015 to December 2020, a total of 1,113 individuals diagnosed with ATAAD and treated surgically at Fuwai Hospital were included in the study. A total of 123 patients who underwent isolated ascending aorta replacement and 147 patients who had ascending aorta with hemi-arch replacement were omitted from the final analysis. 203 chronic Type A aortic dissection patients were also excluded, remaining 640 individuals with TAR+FET. We gathered demographic details, pre-surgery risk elements, and crucial intraoperative data from all patients for examination.

Approval was obtained (no.2020-1402) from the Ethics Committee of Fuwai hospital. The research involved minimal risk to patients, so patient informed consent was not required. The waiver has no negative impact on the participants’ rights and well-being.

### 2.2. Machine learning

In order to compare the effect of different variables on the outcome and identify the key factors, we applied StandardScaler to standardize feature data, transforming the values of each feature into a standard normal distribution with a mean of 0 and a standard deviation of 1 (also known as the Z-score distribution). The variables that met the inclusion criteria were successively entered into each machine learning model, and the AUC test was performed on different models. The predictors were obtained according to the final improved algorithm, and the SHAP was used for interpretation. **Figure 1** illustrates the entire procedure.

**Figure 1.**
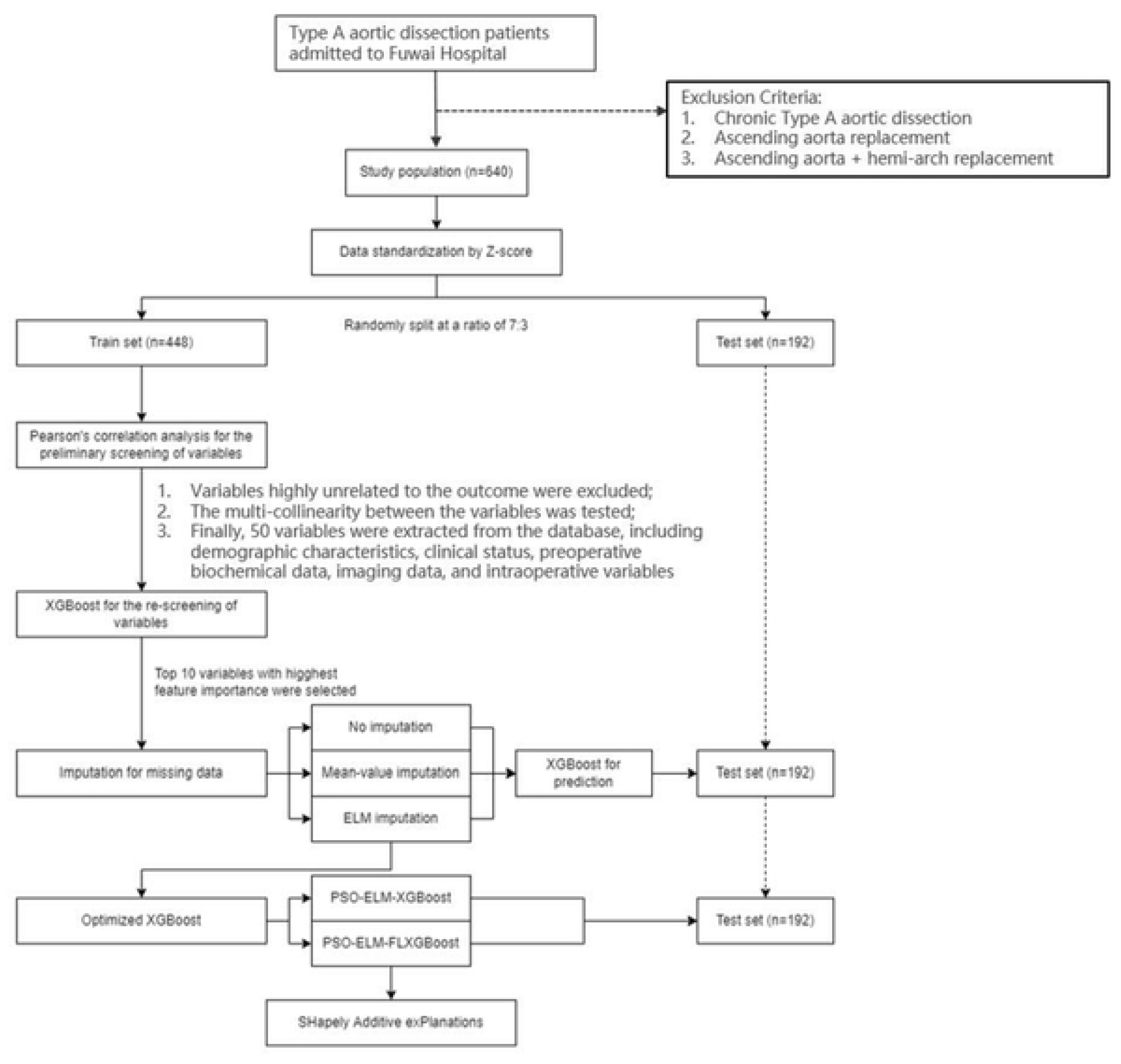
Study Flow chart. Abbreviations: ELM, Extreme Learning Machine; PSO, Particle Swarm Optimization; FL, Focal Loss.

### 2.3. Data Pre-processing

ELM^7^, also known as Extreme Learning Machine, is a type of feed-forward neural network that requires only the specification of the hidden layer’s neuron count. By utilizing random generation of input weights and hidden layer thresholds, acquiring optimal output weights through least squares, and fitting output values through stochastic mapping, this approach offers the benefits of exceptional efficiency and robust generalization capabilities.

### 2.4. Variable selection

Our study chose appropriate variables for model building by multistage screening method. In the first stage, we eliminated variables that significantly unrelated to the outcome. Using Pearson’s correlation analysis, we also examined the multicollinearity between all variables, and selected applicable variables according to the previous research results. In the second stage, we applied XGBoost to generate feature importance of each variable retained in the first stage, indicating their predictive values for the model^8–9^. Ultimately, the top 10 variables with highest feature importance were selected as the predictive factors for the model.

## 3. Model derivation

### 3.1. Prediction model

This study utilized the XGBoost technique and employed the latest ELM to replace the missing data. Additionally, PSO (Particle Swarm Optimization)and FL(Focal Loss) were employed for parameter adjustment, and the model underwent continuous optimization to achieve the best possible machine learning model^10–11^.

### 3.2. Evaluation of prediction model

The validation group was employed to determine the accuracy of different machine learning models by computing the areas under the ROC curve. Moreover, in order to better interpret the association between variables and outcome, our study applied SHapley Additive exPlanation (SHAP) method to assign consistent attribution values to each variable in the model^12^.A variable’s contribution to risk prediction increases as the SHAP absolute value of the variable increases.

### 3.3. Statistical Analysis

All charts were created using the R programming tool (http://www.R-project.org; Version 4.2.1). Statistical significance was determined by a p-value less than 0.05 or an adjusted p-value below 0.05. To compare the differences between the two groups, we conducted a parametric analysis using Student’s t-test and a non-parametric analysis with the Mann-Whitney U test. The p value was adjusted using the Benjamini-Hochberg method, and categorical variables were examined with the Chi-square test. Continuous variables that were normally distributed or nearly normal were represented using the mean and standard deviation. The median [IQR] was used to describe the continuous variables of skewness distribution.

## 4. Results

### 4.1. Training and validation of models

From January 2015 to December 2020, 1113 consecutive patients treated surgically were admitted to the Fuwai Hospital. We selected patients according to the relevant criteria. After normalizing the data with z-scores, the patients were randomly split, assigning 70% to the training group and the other 30% to the testing group. For all variables, the differences between the training set and the test set were nonsignificant. Figure 2 displays the heatmap of the ten most significant variables linked to intensity in the initial stage (using XGBoost exclusively).

**Figure 2.**
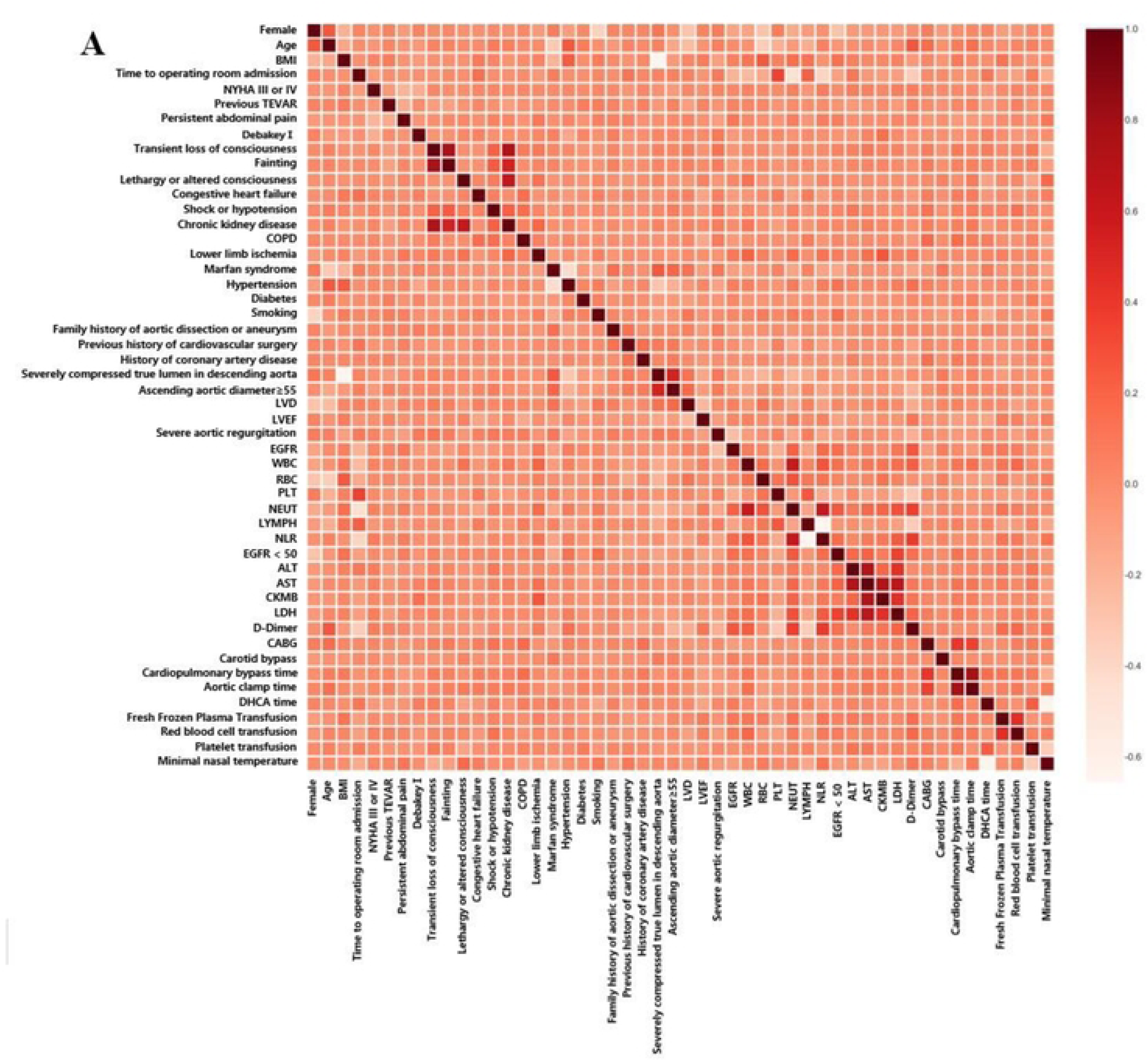
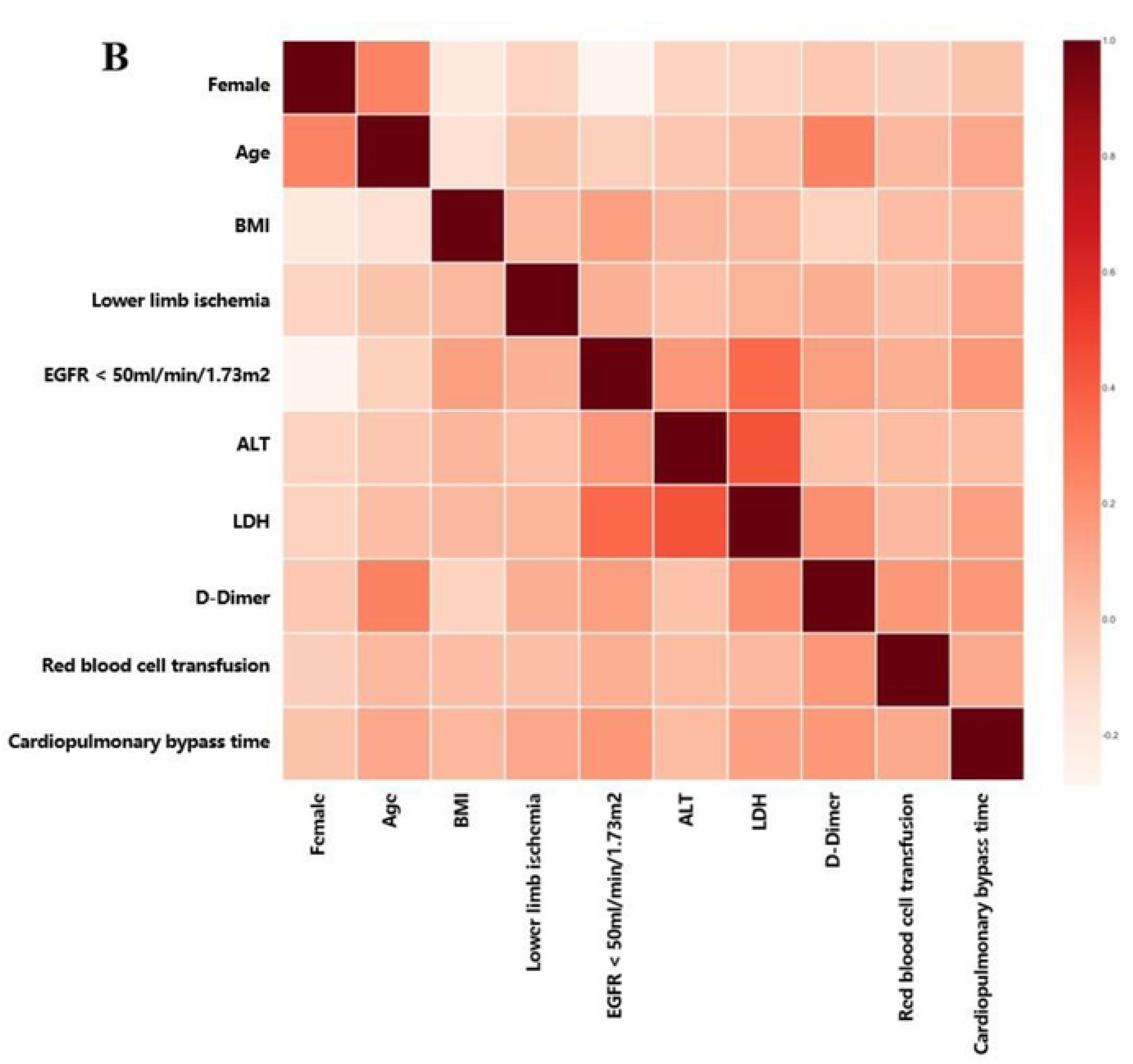
(A) Hot map of the selected variables.(B) The hot map of the top ten variables related to intensity in first stage. ALT, Alanine aminotransferase; AST, Aspartate aminotransferase; BMI, body mass index; CABG, coronary artery bypass grafting; CK-MB, Creatine kinase-MB; COPD, Chronic obstructive pulmonary disease; CPB, cardiopulmonary bypass; EGFR, Estimated Glomerular Filtration Rate; LDH, lactate dehydrogenase; LVD, left ventricular internal diameter; LVEF, left ventricular ejection fraction; LYMPH, lymphocyte; NEUT, neutrophil; NLR, neutrophil to lymphocyte ratio; NYHA, New York Heart Association Classification; PLT, platelets; RBC, red blood cell; TEVAR, thoracic endovascular aortic repair; WBC, white blood cell.

Of all the patients (mean age 46.78±10.12 years, females accounted for 21.09%) enrolled in this study, 83.91% had hypertension, 9.22% had Marfan syndrome. A total of 37 patients (5.78%) developed postoperative 30-Day mortality and were included in the outcome event collection. Table 1 provides a comprehensive overview of the demographic and perioperative data.

**Table 1.**
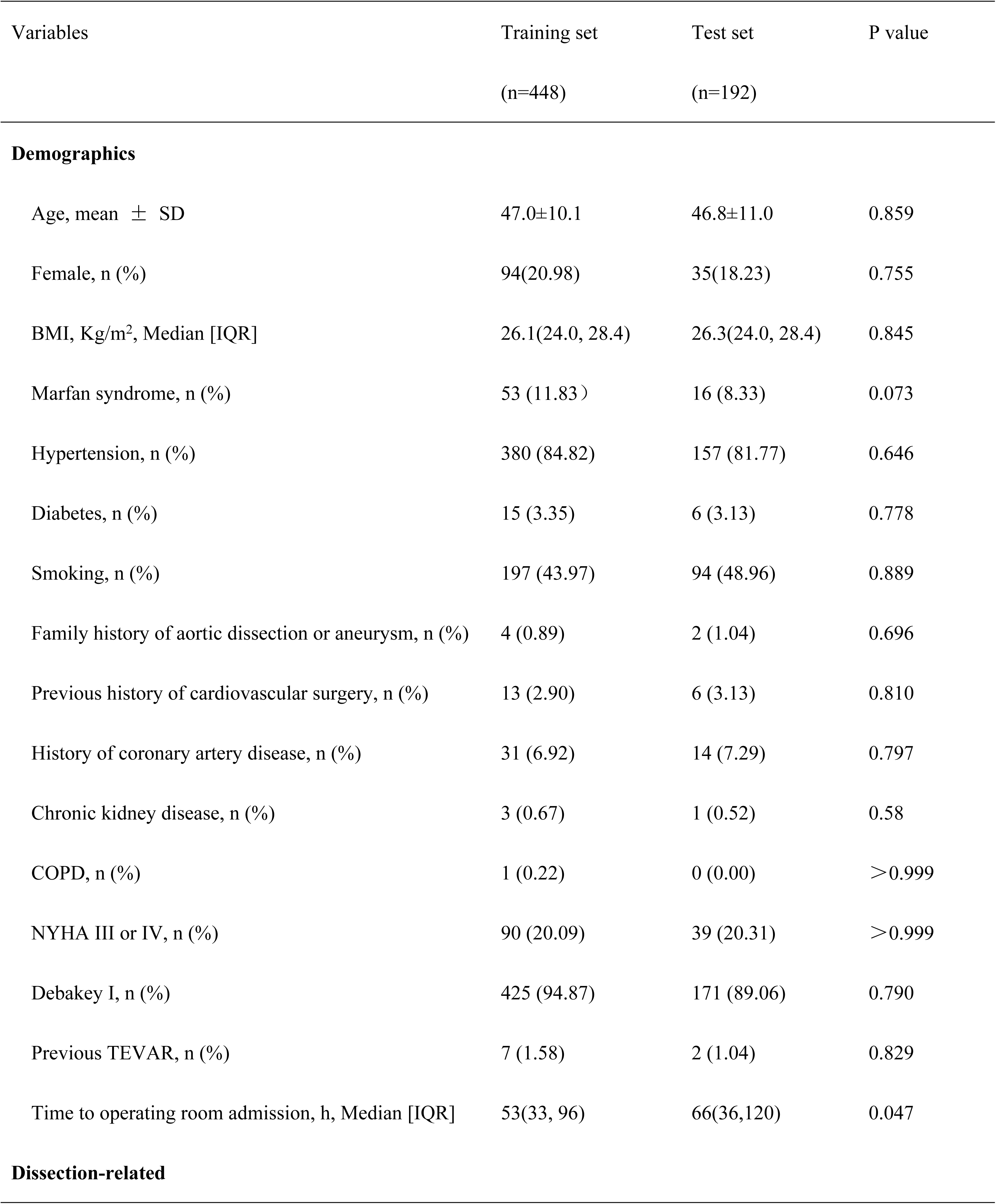
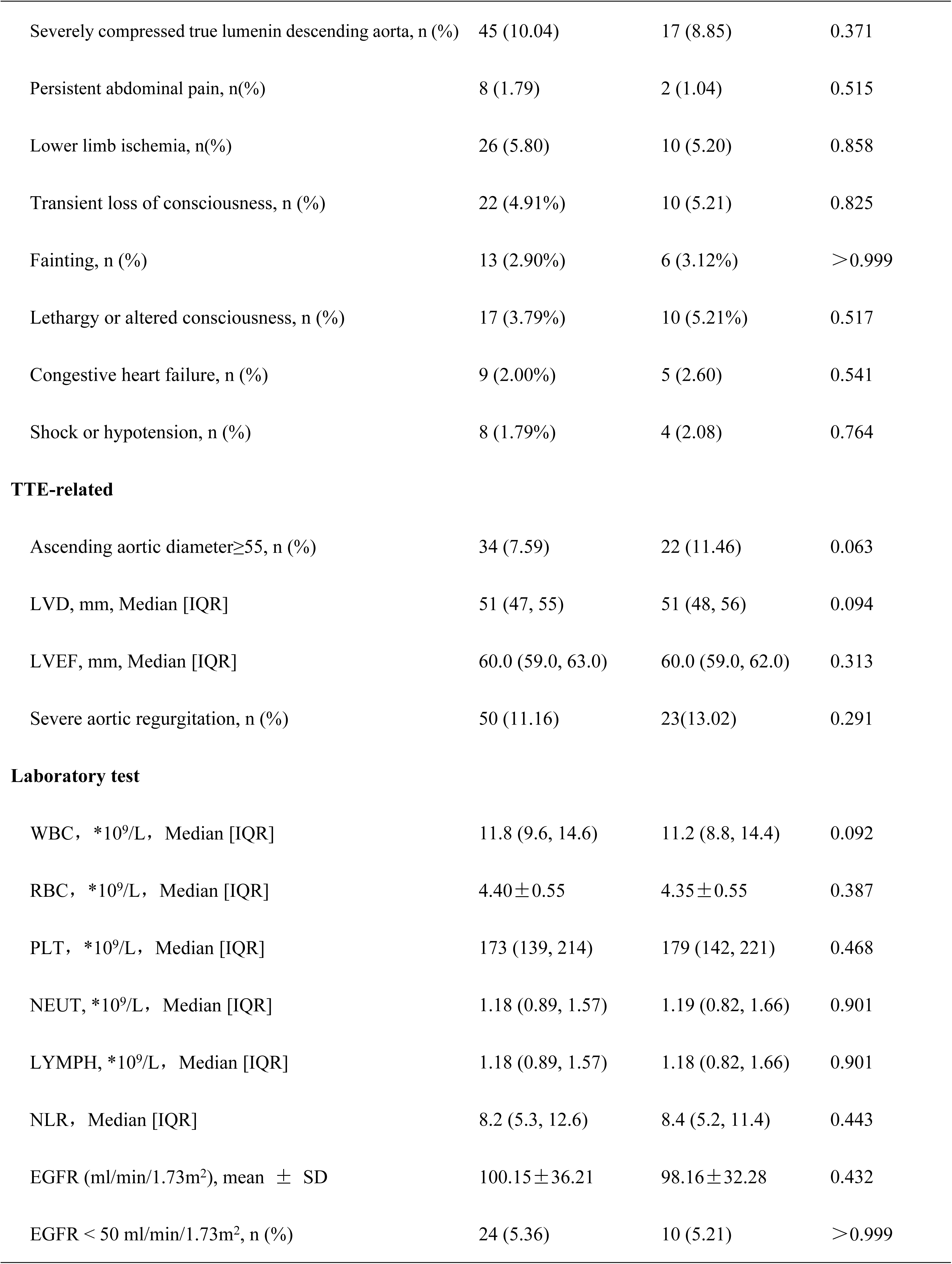
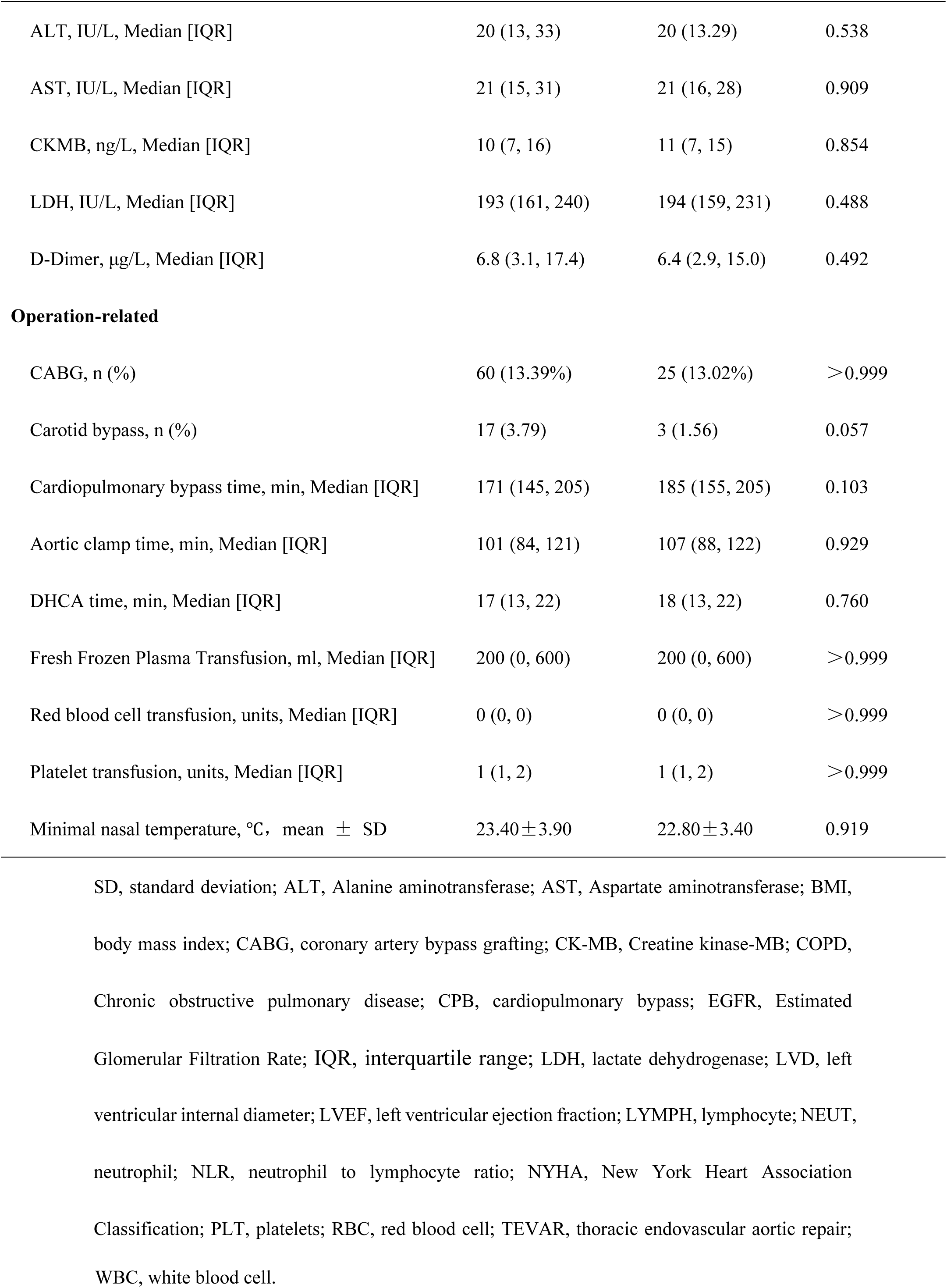
Patient characteristics and perioperative variables are presented in Table 1.

| Variables | Training set<br>(n=448) | Test set<br>(n=192) | P value |
| --- | --- | --- | --- |
| **Demographics** |  |  |  |
| Age, mean ± SD | 47.0±10.1 | 46.8±11.0 | 0.859 |
| Female, n (%) | 94(20.98) | 35(18.23) | 0.755 |
| BMI, Kg/m<sup>2</sup>, Median [IQR] | 26.1(24.0, 28.4) | 26.3(24.0, 28.4) | 0.845 |
| Marfan syndrome, n (%) | 53 (11.83) | 16 (8.33) | 0.073 |
| Hypertension, n (%) | 380 (84.82) | 157 (81.77) | 0.646 |
| Diabetes, n (%) | 15 (3.35) | 6 (3.13) | 0.778 |
| Smoking, n (%) | 197 (43.97) | 94 (48.96) | 0.889 |
| Family history of aortic dissection or aneurysm, n (%) | 4 (0.89) | 2 (1.04) | 0.696 |
| Previous history of cardiovascular surgery, n (%) | 13 (2.90) | 6 (3.13) | 0.810 |
| History of coronary artery disease, n (%) | 31 (6.92) | 14 (7.29) | 0.797 |
| Chronic kidney disease, n (%) | 3 (0.67) | 1 (0.52) | 0.58 |
| COPD, n (%) | 1 (0.22) | 0 (0.00) | >0.999 |
| NYHA III or IV, n (%) | 90 (20.09) | 39 (20.31) | >0.999 |
| Debaquey I, n (%) | 425 (94.87) | 171 (89.06) | 0.790 |
| Previous TEVAR, n (%) | 7 (1.58) | 2 (1.04) | 0.829 |
| Time to operating room admission, h, Median [IQR] | 53(33, 96) | 66(36,120) | 0.047 |
| **Dissection-related** |  |  |  |
| Severely compressed true lumen in descending aorta, n (%) | 45 (10.04) | 17 (8.85) | 0.371 |
| Persistent abdominal pain, n(%) | 8 (1.79) | 2 (1.04) | 0.515 |
| Lower limb ischemia, n(%) | 26 (5.80) | 10 (5.20) | 0.858 |
| Transient loss of consciousness, n (%) | 22 (4.91%) | 10 (5.21) | 0.825 |
| Fainting, n (%) | 13 (2.90%) | 6 (3.12%) | >0.999 |
| Lethargy or altered consciousness, n (%) | 17 (3.79%) | 10 (5.21%) | 0.517 |
| Congestive heart failure, n (%) | 9 (2.00%) | 5 (2.60) | 0.541 |
| Shock or hypotension, n (%) | 8 (1.79%) | 4 (2.08) | 0.764 |
| **TTE-related** |  |  |  |
| Ascending aortic diameter $\geq 55$, n (%) | 34 (7.59) | 22 (11.46) | 0.063 |
| LVD, mm, Median [IQR] | 51 (47, 55) | 51 (48, 56) | 0.094 |
| LVEF, mm, Median [IQR] | 60.0 (59.0, 63.0) | 60.0 (59.0, 62.0) | 0.313 |
| Severe aortic regurgitation, n (%) | 50 (11.16) | 23 (13.02) | 0.291 |
| **Laboratory test** |  |  |  |
| WBC, $\times 10^9/L$, Median [IQR] | 11.8 (9.6, 14.6) | 11.2 (8.8, 14.4) | 0.092 |
| RBC, $\times 10^9/L$, Median [IQR] | 4.40 ± 0.55 | 4.35 ± 0.55 | 0.387 |
| PLT, $\times 10^9/L$, Median [IQR] | 173 (139, 214) | 179 (142, 221) | 0.468 |
| NEUT, $\times 10^9/L$, Median [IQR] | 1.18 (0.89, 1.57) | 1.19 (0.82, 1.66) | 0.901 |
| LYMPH, $\times 10^9/L$, Median [IQR] | 1.18 (0.89, 1.57) | 1.18 (0.82, 1.66) | 0.901 |
| NLR, Median [IQR] | 8.2 (5.3, 12.6) | 8.4 (5.2, 11.4) | 0.443 |
| EGFR (ml/min/1.73m<sup>2</sup>), mean ± SD | 100.15 ± 36.21 | 98.16 ± 32.28 | 0.432 |
| EGFR < 50 ml/min/1.73m<sup>2</sup>, n (%) | 24 (5.36) | 10 (5.21) | >0.999 |
| ALT, IU/L, Median [IQR] | 20 (13, 33) | 20 (13.29) | 0.538 |
| AST, IU/L, Median [IQR] | 21 (15, 31) | 21 (16, 28) | 0.909 |
| CKMB, ng/L, Median [IQR] | 10 (7, 16) | 11 (7, 15) | 0.854 |
| LDH, IU/L, Median [IQR] | 193 (161, 240) | 194 (159, 231) | 0.488 |
| D-Dimer, µg/L, Median [IQR] | 6.8 (3.1, 17.4) | 6.4 (2.9, 15.0) | 0.492 |
| **Operation-related** |  |  |  |
| CABG, n (%) | 60 (13.39%) | 25 (13.02%) | >0.999 |
| Carotid bypass, n (%) | 17 (3.79) | 3 (1.56) | 0.057 |
| Cardiopulmonary bypass time, min, Median [IQR] | 171 (145, 205) | 185 (155, 205) | 0.103 |
| Aortic clamp time, min, Median [IQR] | 101 (84, 121) | 107 (88, 122) | 0.929 |
| DHCA time, min, Median [IQR] | 17 (13, 22) | 18 (13, 22) | 0.760 |
| Fresh Frozen Plasma Transfusion, ml, Median [IQR] | 200 (0, 600) | 200 (0, 600) | >0.999 |
| Red blood cell transfusion, units, Median [IQR] | 0 (0, 0) | 0 (0, 0) | >0.999 |
| Platelet transfusion, units, Median [IQR] | 1 (1, 2) | 1 (1, 2) | >0.999 |
| Minimal nasal temperature, °C, mean ± SD | 23.40±3.90 | 22.80±3.40 | 0.919 |

SD, standard deviation; ALT, Alanine aminotransferase; AST, Aspartate aminotransferase; BMI,
body mass index; CABG, coronary artery bypass grafting; CK-MB, Creatine kinase-MB; COPD,
Chronic obstructive pulmonary disease; CPB, cardiopulmonary bypass; EGFR, Estimated
Glomerular Filtration Rate; IQR, interquartile range; LDH, lactate dehydrogenase; LVD, left
ventricular internal diameter; LVEF, left ventricular ejection fraction; LYMPH, lymphocyte; NEUT,
neutrophil; NLR, neutrophil to lymphocyte ratio; NYHA, New York Heart Association
Classification; PLT, platelets; RBC, red blood cell; TEVAR, thoracic endovascular aortic repair;
WBC, white blood cell.

### 4.2. Model Evaluation and Interpretation

Figure 3 displays the AUCs obtained by employing the enhanced PSO-ELM-FLXGBoost machine learning technique, utilizing all variables as input variables. The AUC (0.869) for the improved model was largest.

**Figure 3.**
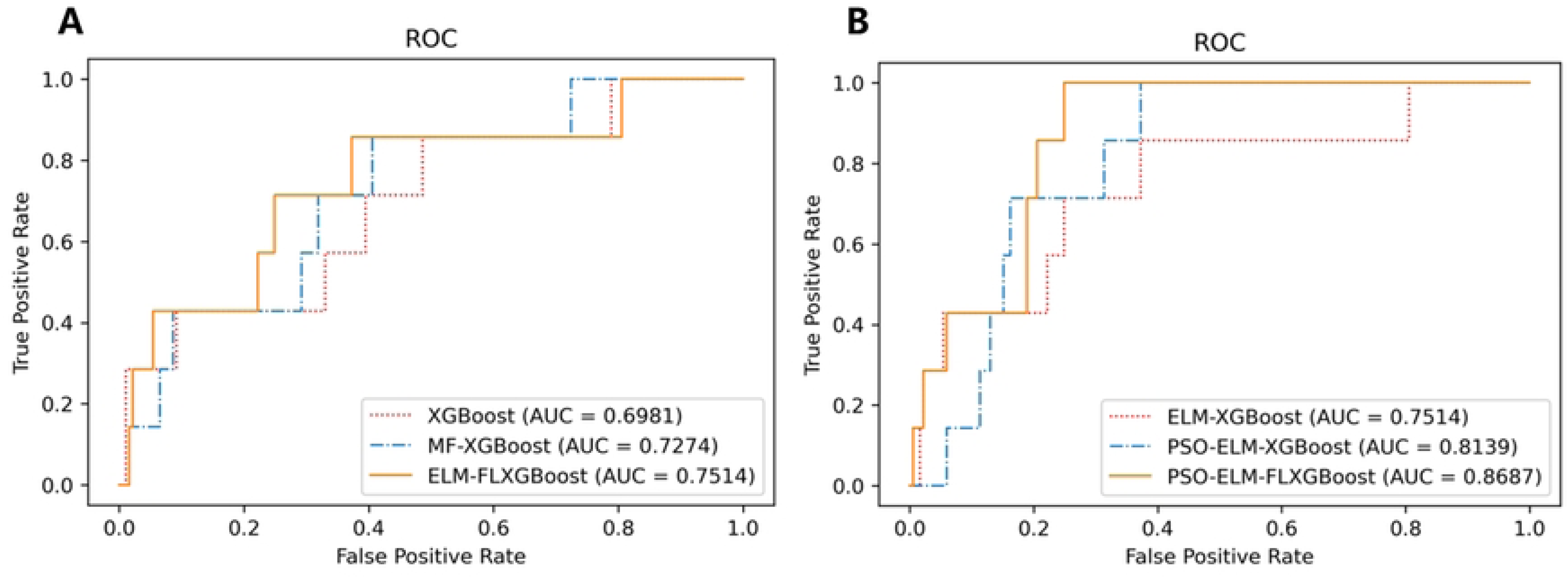
By implementing new methods, the AUC value significantly improved from 0.6981 using the XGBoost model to 0.8687 with the PSO-ELM-FLXGBoost model. (A) The AUC (0.7514) for the ELM-FLXGBoost model was larger than that for the MF-XGBoost (0.7274) and XGBoost (0.6981). (B)The AUC (0.8687) for the PSO-ELM-FLXGBoost model was largest.

Figure 4 displays the significance matrix chart for the PSO-FLXGBoost technique, indicating that the model’s top 10 influential factors include Gender, age, BMI, Lower limb ischemia, EGFR < 50 ml/min/1.73m^2^, ALT, LDH, D-Dimer, transfusion of red blood cells and Cardiopulmonary bypass time.

**Figure 4.**
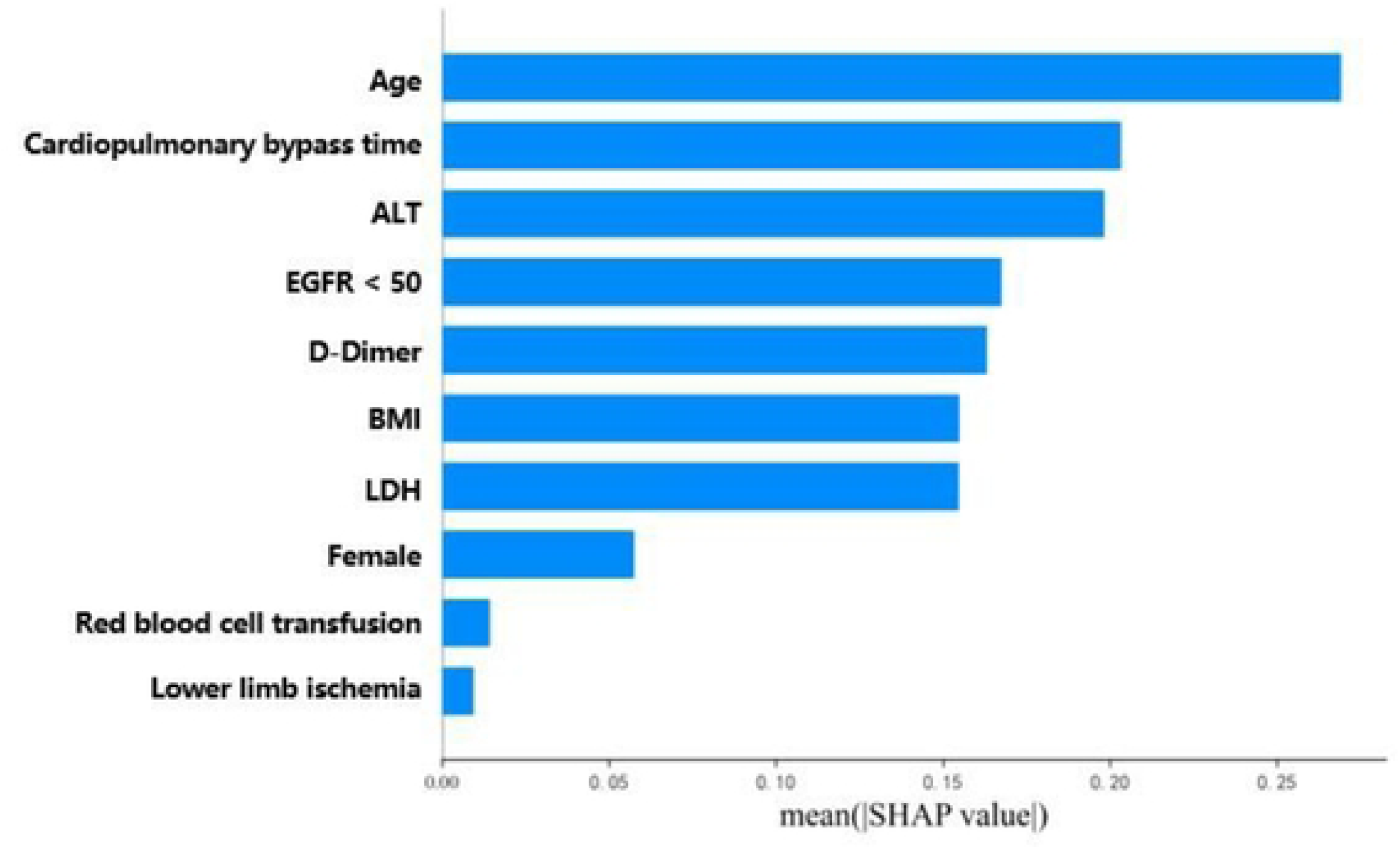
Plotting the matrix of importance for the PSO-FLXGBoost technique. The model is influenced by the top 10 variables that are of utmost significance.

The SHAP summary plot was utilized to determine the characteristics that had the greatest impact on the prediction model. We combine the feature value of the y-axis and the SHAP value of the x-axis, we can see that older age, longer CPB time, higher ALT, D-Dimer and EGFR can significantly increase the probability of perioperative mortality. SHAP analysis of all significant variables is shown in Figure 5.

**Figure 5.**
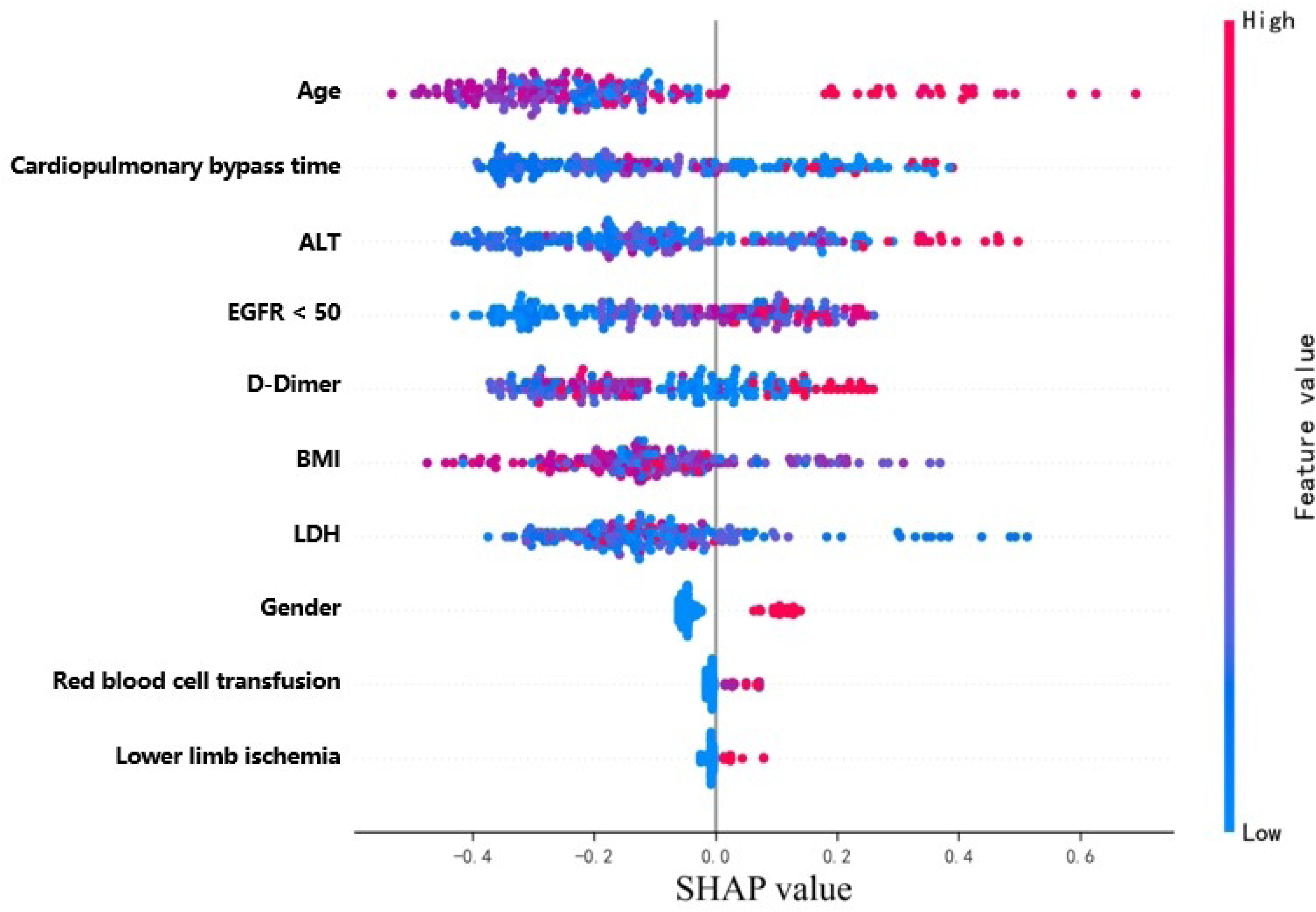
SHAP analysis of top 10 significant variables.

## 5. Discussion

According to data from the International Registry of Aortic Dissection (IRAD) cohort, the mean age of patients is 61.5 ± 14.6 years^13^. However, in China, the mean age of patients with ATAAD is significantly younger at 46.8 ± 12.1 years, which may be due to differences in aortic pathology and poor blood pressure control, resulting in a higher proportion of Chinese TAAA patients with Marfan syndrome and aortic dissection. Furthermore, a retrospective study of ATAAD patients revealed that TAR+FET is frequently employed in patients under 50 years of age^14^. This surgical approach is preferred by surgeons due to the longer life expectancy of Chinese patients.

Despite the relative standardization of the TAR procedure and advancements in perioperative care, postoperative mortality following surgical intervention remains high. Accurate assessment of the condition’s severity and timely intervention can significantly enhance patient outcomes and reduce mortality rates. Thus, the development of effective predictive modeling is essential to evaluate associated risks and identify higher-risk populations within clinical settings.

In our past work, we used a traditional logistic way to develop and validate a nomogram model and applied it in clinical situations^15^. The nomogram is a simple and effective tool in our clinical practice. However, no method is perfect. Regression model explain well but are not flexible enough, and machine learning methods are flexible but have limited explanatory power. Clinical predictive modeling as an intelligent tool for risk and benefit assessment, how we optimize the model is an important issue.

With the development of machine learning predictive models, we note that ML leverages artificial intelligence to autonomously extract valuable information, identify potential patterns within large datasets, and develop robust risk models^16^. Compared to traditional analytical methods, ML demonstrates superior predictive strength and stability while addressing nonlinear and complex interactions between variables and outcomes. As a result, ML is increasingly employed across various medical domains, including diagnosis, prognostic prediction, treatment, and medical image analysis^17^. However, there is a notable lack of research specifically focused on the use of machine learning to evaluate the risk of perioperative mortality in individuals diagnosed with Acute Type A Aortic Dissection following TAR+FET.

It is widely recognized that machine learning techniques are often used on extensive datasets. While the sample size in this research is adequate for TAR+FET, it is somewhat limited for machine learning algorithms, potentially leading to subpar model performance. To optimize the prediction, Particle Swarm Optimization (PSO), an intelligent algorithm that simulates bird behavior, was employed. Additionally, considering the small proportion of individuals with adverse outcomes, we employed Imbalance-XGBoost to improve the predictions. This involved applying PSO and the FL function to further enhance the algorithm.

To better interpret the association between variables and outcomes, our study employed the SHAP (SHapley Additive exPlanations) method, which assigns consistent attribution values to each variable in the model^18^. Variables with higher SHAP absolute values made greater contributions to risk prediction. The PSO-ELM-FLXGBoost model outperformed the other models in terms of prediction, achieving the highest AUC among all the models tested. Moreover, this model demonstrated improved predictive power even with small datasets or imbalanced outcome events compared to the total sample size. We hope that this optimized PSO-ELM-FLXGBoost model can inform the construction of machine learning in clinical prediction models.

In our study, machine learning methods were successfully established to predict ATAAD perioperative mortality in our cohort. But most importantly, The ultimate goal of medical predictive model construction is for clinical application. We hope to provide some ideas for future predictive modeling-assisted clinical decision making and believe that traditional regression modeling and machine learning approaches each have their own characteristics and can be better applied to clinical practice if they complement each other. We are looking forward to optimizing the model in the follow-up study to combine the traditional model with the machine learning approach for better clinical applications.

## 6. Limitations

There are various constraints in this study that require attention. It is crucial to mention that this study is retrospective and conducted in a single center, which could lead to bias because of patients at a single center may not be broadly representative of the general population. Additionally, the effectiveness of machine learning algorithms may differ based on the dataset’s magnitude and the distribution of patient attributes. Moreover, it is crucial to acknowledge that this model lacks external validation, potentially restricting its applicability and reducing its accuracy in representing real-life situations. Hence, additional research is required to assess the feasibility of this model.

## 7. Conclusions

To sum up, this research employed preoperative lab tests, clinical imaging records, and initial postoperative therapy results of individuals diagnosed with aortic dissection. Our team has effectively created and verified machine learning algorithms for forecasting the occurrence of death within 30 days after surgery in individuals diagnosed with acute type A aortic dissection. The death risk model suggested in this research is simple and easy to use, allowing medical practitioners to evaluate the risks associated with the condition and treatment, anticipate the probability of mortality in individuals with aortic dissection, direct the creation and execution of suitable and efficient treatment strategies, improve prognosis and survival rates, and ultimately decrease the rates of mortality.

## Data Availability

Data cannot be shared publicly because of data sharing agreements and research ethics board protocols with participating hospitals.. Data are available from the Fuwai hospital Ethics Committee (contact via https://www.fuwai.com/News/Articles/Index/192) for researchers who meet the criteria for access to confidential data.

